# Identification and application of plasmatic microRNA expression quantitative trait loci (miR-QTL) at first trimester of pregnancy

**DOI:** 10.1101/2021.11.30.21267083

**Authors:** Frédérique White, Marika Groleau, Samuel Côté, Cécilia Légaré, Kathrine Thibeault, Andrée-Anne Clément, Marie-France Hivert, Luigi Bouchard, Pierre-Étienne Jacques

## Abstract

**Background:** MicroRNAs (miRNAs) are a class of small non-coding RNAs regulating gene expression. They are involved in many biological processes, including adaptation to pregnancy. The identification of genetic variants associated with gene expression, known as expression quantitative trait loci (eQTL), helps to understand the underlying molecular mechanisms and determinants of complex diseases. Using data from the prospective pre-birth Gen3G cohort, we investigated associations between maternal genotypes and plasmatic miRNA levels measured during the first trimester of pregnancy of 369 women.

**Results:** Assessing the associations between about 2 million SNPs and miRNA proximal pairs using best practices from the GTEx consortium, a total of 22,140 significant eQTLs involving 147 unique miRNAs were identified. Elastic-net regressions were applied to select the most relevant SNPs to build genetic risk scores (GRS) for each of these 147 miRNAs. For about half of the circulating miRNAs, the GRS captured >10% of the variance abundance. As a demonstration of the usefulness of the identified eQTLs and derived GRS, we used the GRSs as instrumental variables to test for association between the circulating levels of miRNAs quantified before the 16th week of pregnancy and the development of pregnancy complications (gestational diabetes [GDM] or pre-eclampsia [PE]) developing more than three months later on average. Using predicted miRNA levels derived from instrumental variables, we found 18 significant associations of miRNAs with potential support of causal inference for GDM or PE.

**Conclusions:** Our results represent a valuable resource to understand miRNA regulation and highlight the potential of genetic instruments in predicting circulating miRNA levels and their possible contribution in disease development.

## Background

MicroRNAs (miRNAs) are small non-coding RNAs regulating gene expression by gene silencing effects [1], with approximately 60% of human mRNA being conserved targets of miRNAs [2]. miRNAs are therefore involved in many biological processes and conditions. Circulating miRNAs in serum and plasma are secreted by different cell types. These miRNAs are known for their diversity, abundance and stability [3], but understanding their own regulation still requires effort [4].

The statistical association of single nucleotide polymorphisms (SNPs) with gene expression, known as expression quantitative trait loci (eQTL), helps to understand the underlying molecular mechanisms of gene regulation and determinants of complex diseases [5]. Although circulating miRNAs have been suggested to be promising biomarkers, the capacity to predict their levels would be of great interest as their quantification remains technically challenging. Genetic Risk scores (GRSs) are used to estimate the genetic contribution to a phenotype of interest. GRSs can then be used to predict a medical condition (when used in context of precision medicine), or a biomarker such as the plasmatic miRNA level to use in causal inferences analyses.

Using data from the prospective pre-birth cohort Genetics of Glucose Regulation in Gestation and Growth (Gen3G), we investigated associations between maternal genotypes and miRNA levels in plasma during the first trimester of pregnancy. For this analysis, we used a subset of 369 samples from which maternal genotype and plasma miRNA quantification data were both available. We report genome-wide proximal miRNA-eQTLs (*cis* miR-QTLs) to provide insight into the regulation of miRNAs in the context of pregnancy. In addition, knowledge of the regulation of miRNAs during pregnancy could provide additional information on pregnancy-associated diseases.

## Results

### Genotypic and microtranscriptomic data processing

In the Gen3G prospective pre-birth cohort built to investigate glucose regulation determinants in pregnancy and its impact on fetal development and children’s health [6], the genotyping data was available for 569 self-reported European ancestry mothers. Levels of plasmatic miRNA in the first trimester of pregnancy were available for a subset of 369 of these mothers (Table S1). After imputation and filtering of the genotyping data, ∼5.4 million autosomal SNPs with minor allele frequency (MAF) >0.05, imputation accuracy >0.8 and Hardy-Weinberg equilibrium (HWE) *p*-value >1e-8 were selected for the current study (Fig. 1A). We conducted a PCA to detect population substructure and confirm the reported European ancestry of the participants by jointly analyzing the data from Gen3G and the 1000 Genomes Project (Fig. S1A-D). Quantification of the microtranscriptomic data was performed with the exceRpt pipeline [7] using miRBase v21 annotations [8] (Fig. 1B). The 1744 detected miRNAs originating from a single genomic loci were filtered to select 550 miRNAs minimally expressed in at least 20% of the samples, similar to filters applied previously [9].

**Figure 1.**
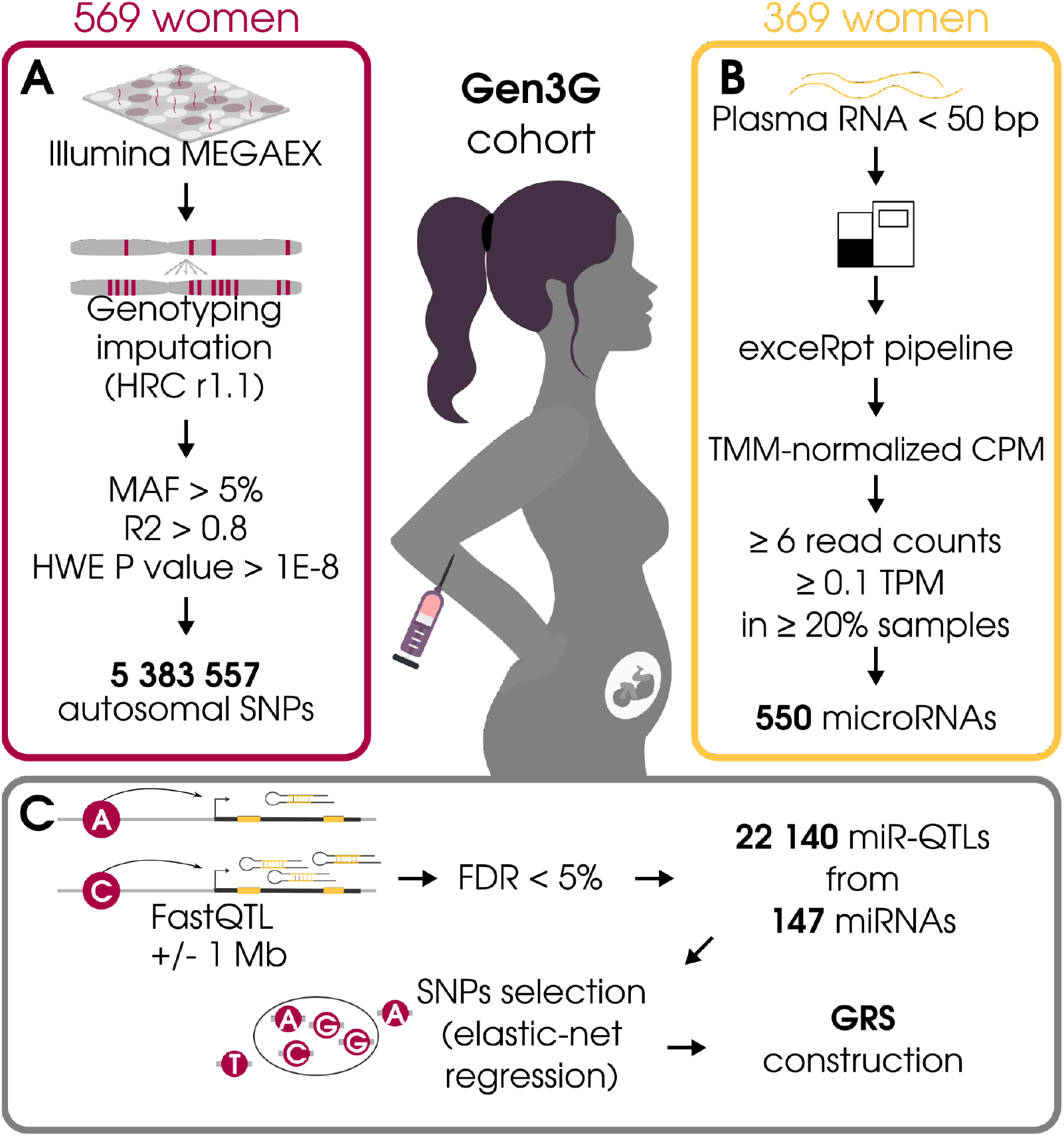
Schematic overview of miR-QTL identification and application from the Gen3G cohort. A) Processing of the genotyping data. B) Processing of the miRNA expression data. C) miR-QTL identification and application

### miR-QTL identification and characterization

Based on best practices outlined by GTEx [9], we investigated the associations between the selected ∼5.4M SNPs and the measured levels of the selected 550 circulating plasma miRNAs. Using FastQTL [10], we tested 2,130,907 *cis-*SNP-miRNA pairs (Fig. 1C, Fig. S1E) and found 22,140 significant miR-QTLs associated with the abundance of 147 miRNAs (FDR < 0.05) (Fig. 2A, Table S2). As expected based on their low proportion of expressed miR, no associations were found on chromosome 12 and 18.

**Figure 2.**
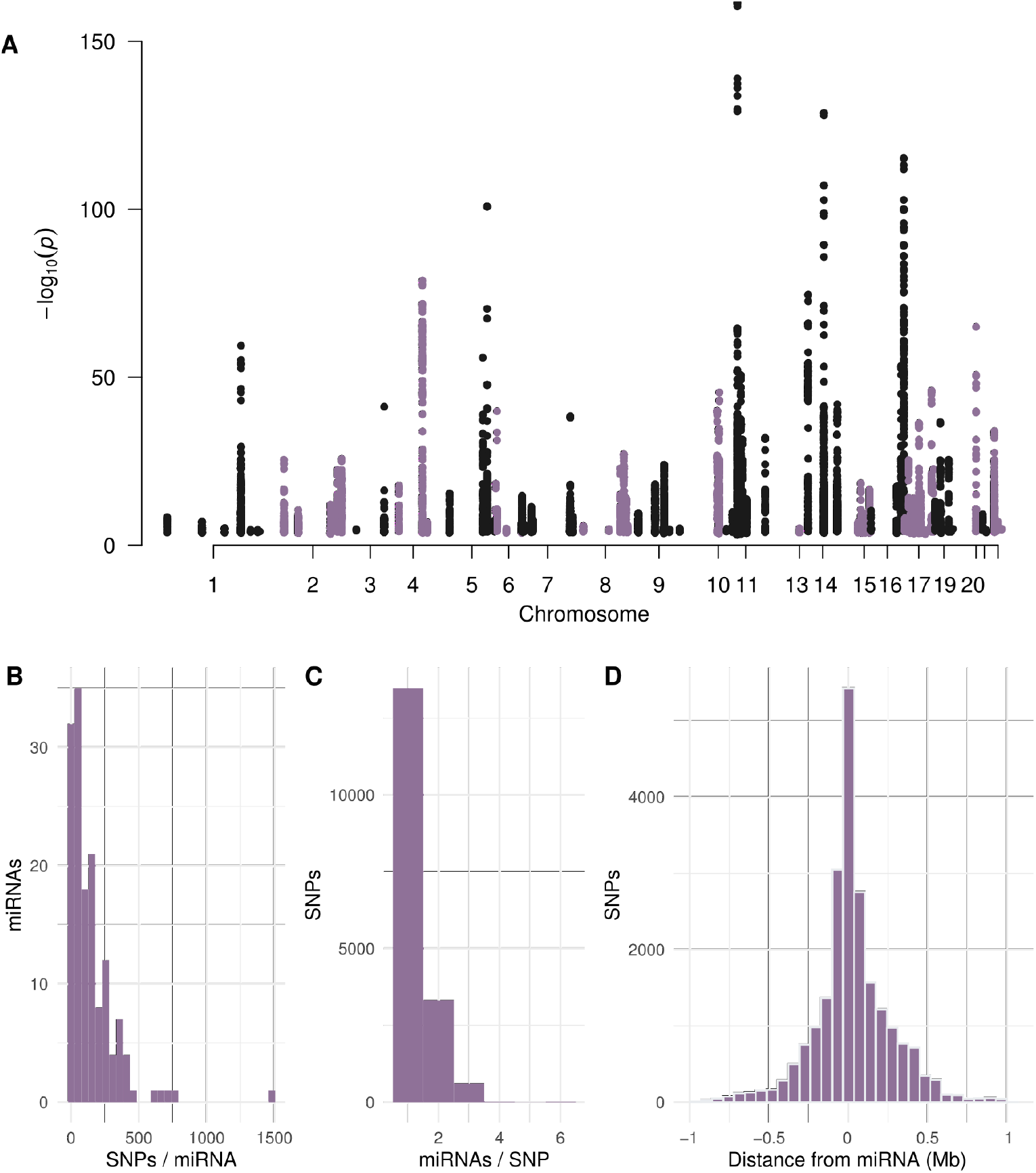
Characterization of the miR-QTLs. A) Manhattan plots representing the genome-wide distribution of the miR-QTLs. B) Distribution of the number of SNP significantly associated per miRNA. C) Distribution of the number of miRNA significantly associated per SNP. D) Distribution of relative distance between the SNPs and the associated miRNAs.

Individual miRNAs were associated between 1 to 1481 SNPs (mean=151, median = 104) (Fig. 2B). The most significant *cis-*SNP-miRNA associations also showed the highest number of associated SNPs (hsa-miR-3161, Fig. S2). The vast majority of the SNPs (77%) were associated with only one miRNA (Fig. 2C). Interestingly, all those associated with more than three miRNAs were located in the known miRNA cluster on chromosome 14 (C14MC). About a third of the miR-QTL were within 50 kb of their corresponding mature miRNA (Fig. 2D, Table S1).

### Application of the miR-QTL

In addition to the informative individual miR-QTL associations, we created genetic risk scores (GRS) for each of the 147 miRNAs identified with cis-QTL to summarize the effect of multiple miR-QTLs on their plasma abundance. We applied elastic-net regressions to select the most relevant SNPs among the miR-QTLs identified and then computed their weight in the GRS. Most of the GRSs (∼80%) contained less than 30 SNPs and about half of them (n=69 GRSs, 47%) captured more than 10% of the variance of their associated miRNA (Figure 3, Table S2-3).

**Figure 3.**
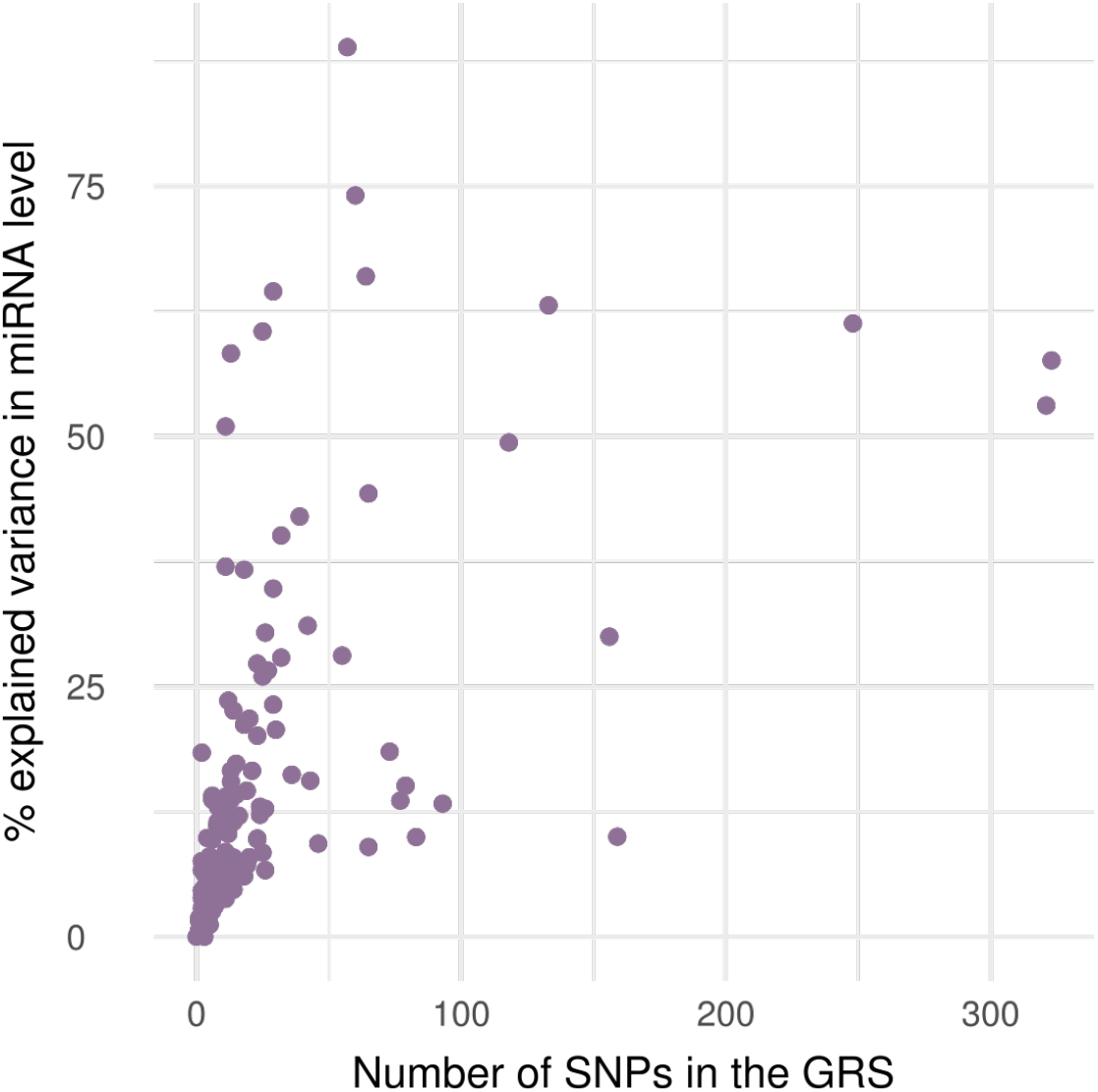
Distribution of the number of SNPs in each GRS and their proportion of miRNA abundance variance explained.

One of the very interesting features of the GRS is the ability to use it as an instrumental variable in causal inference analyses to predict a phenotype or biomarker (in this case, the circulating miRNA levels) solely based on the genotype. To demonstrate the potential of such instruments for circulating plasma miRNAs in the context of pregnancy, we used the GRSs to test the association between predicted miRNA levels and the three most frequent pregnancy complications (gestational diabetes [GDM], preeclampsia [PE] and gestational hypertension (GH)). Regressing observed miRNA levels on GRSs, we estimated the abundance of each of the 147 miRNA for all of the mothers for which we had the genotype, thus adding 200 samples for which plasma miRNA levels data were missing. We then used logistic regressions to evaluate the association between the predicted miRNA abundance and each outcome and found 8 miRNAs associated with GDM (p-value < 0.05), including hsa-miR-323b-3p that belongs to the chromosome 14 microRNA cluster (C14MC) (Table 1). In this case, there were more than 3000 SNPs genotyped with a MAF>5% within the selected +/- 1Mb window, from which 76 were identified as eQTLs and 21 were selected for its GRS, overall explaining ∼17% of variance in hsa-miR-323b-3p levels (Fig. S3A). In the regression analysis using the measured plasma miRNA level in 369 women, higher levels of hsa-miR-323b-3p in early pregnancy was associated with lower risk of GDM (n=41) although this association was not significant (observed OR= 0.74; 95%CI= 0.53-1.04; Fig. 4A, left). Using the 21-SNPs GRS as an instrumental variable (total n= 569, including n= 49 GDM cases), higher predicted levels of hsa-miR-323b-3p were significantly associated with lower risk of GDM (OR= 0.45; 95%CI= 0.23-0.92; Fig. 4A, middle and right). Equivalent analyses were conducted for the two other complications and identified 5 miRNAs associated with PE and 3 with GH (Table 1). We are presenting examples for PE using hsa-miR-873-3p and for GH using hsa-miR-4772-5p (Fig. 4B-C).

**Figure 4.**
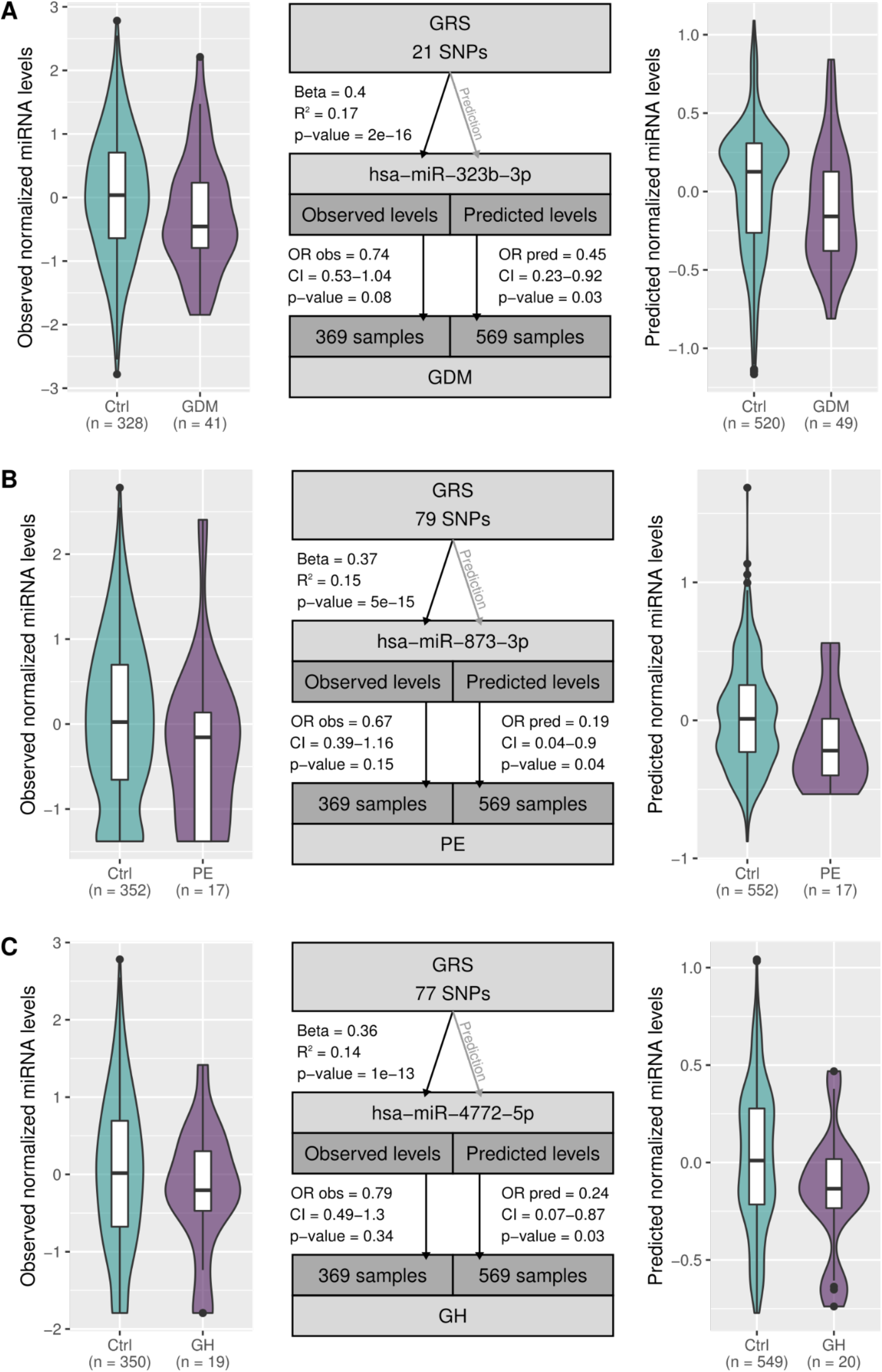
Examples of GRSs as an instrumental variable for miRNA expression and association with three pregnancy complications reported in Gen3G. The complications are GDM (A), PE (B) and GH (C). Instrumental variables usage are presented in the middle. On each side, the distributions of observed (left) and predicted (right) miRNA levels are shown for cases and controls over the 369 and 569 samples, respectively.

## Discussion

We aimed at mapping *cis-*QTL for plasmatic miRNAs in the context of pregnancy. The regulation of miRNA expression and abundance is still poorly understood [11], and the identification of miR-QTLs addresses this issue by highlighting potentially important regulating regions of miRNAs. This approach has been used extensively for mRNAs in several tissues, namely by the GTEx consortium, but only few studies have similarly analyzed together microtranscriptomic and genetic data [12–18], and none in the context of pregnancy.

Even though the mapping window for proximal miR-QTL discovery was set to 2Mb, about a third of the miR-QTL were within 50Kb of their corresponding mature miRNA, which is somehow expected for proximal transcriptional regulators. Whether these miR-QTL are within known regulatory regions for miRNA should be investigated in future analyzes, for instance by comparing our results to the FANTOM5 database [19] and other epigenomic resources such as EpiMap, a compendium comprising 10,000 epigenomic maps across 800 samples used to define chromatin states, high-resolution enhancers, enhancer modules [20]. For the miRNAs that are located inside protein-coding or non-coding genes, comparing our miR-QTL to their corresponding eQTLs identified by GTEx could help distinguish between the variability in miRNA abundance that is attributable to host-gene expression and those that are more likely due to RNA processing [9].

Another interesting challenge is the identification of miR-QTL for miRNAs that are located in imprinted clusters, i.e. expressed only from one parental allele. For example, we identified miR-QTL for hsa-miR-323b-3p, which is located in the paternally-imprinted C14MC, a miRNA cluster of interest in the context of pregnancy given its strong expression by the placenta [21]. Another study also previously identified miR-QTLs for this miRNA, although in non-pregnant individuals [18]. Because of the random nature of allele distribution at conception, heterozygous SNPs could not possibly be informative for miR-QTL in imprinted regions without phasing. This means that we likely detected miR-QTLs in imprinted regions based on the information content of homozygous individuals which, because of the consequently reduced power, implies a strong effect of those SNPs on the miRNA abundance. Further analyzes are needed to test this hypothesis.

In about half of the circulating miRNAs with at least 1 cis-QTL, the GRS we derived from the miR-QTLs captured more than 10% of the variance of their respective circulating miRNA, which is overall impressive when considering the many steps where other regulatory mechanisms can act between their nuclear expression and the cirulating levels in plasma. Our results support the high importance of genetic variants in miRNA expression regulation. We were fortunate to have an additional 200 individuals with genotypes available to study the association of predicted plasma miRNAs with the most frequent pregnancy complications, GDM, PE and HG using GRSs as instrumental variables. Interestingly, 15 out of the 18 significant associations identified using the predicted miRNA levels were not significant (p-value > 0.05) when the same associations were relying on the measured miRNA levels, most probably because we increased our sample size. Other reasons could be that the ‘observed’ associations are negatively confounded by other variables not included in the regression analyses, suggesting that the ‘causal’ pathways are stronger than the simple ‘observational’ analyses appear.

## Conclusions

In this study, we identified 22,140 miR-QTLs associated with 147 plasmatic miRNAs detectable in the first trimester of pregnancy. These miR-QTLs are informative of genetic variants likely regulating expression of miRNAs. By combining multiple miR-QTLs into GRSs, we were able to explain substantial degree of plasmatic miRNA level variability. Overall, our results highlight the potential of genetic instruments in predicting circulating miRNA levels and their association with complex traits. Such instruments could help to understand the regulation of miRNA expression and the etiology of complex traits such as pregnancy complications.

## Methods

### Cohort description

For this study, we included 569 women of European ancestry recruited in the Genetics of Glucose regulation in Gestation and Growth (Gen3G). Gen3G is a prospective pre-birth cohort built to investigate glucose regulation determinants in pregnancy and its impact on fetal development and children’s health [6] (Table S1). Twelve non-European samples were excluded as the sample size of the otherwise genetically homogeneous cohort did not allow to adequately leverage a greater genetic diversity.

### Genotype data processing

The genotype data was processed as described in Hivert et al. 2020 [22]. Briefly, genotyping was performed with the Illumina Expanded Multi-Ethnic Genotyping Array (MEGAEx), pre-phased using ShapeIT v2.r790, then imputed using the Haplotype Reference Consortium (HRC) r1.1 reference panel with Minimac3 from the Michigan Imputation Server [23]. From the 39,183,141 resulting autosomal SNPs, we excluded SNPs with MAF ≤0.05, R² score (imputation estimated accuracy) <0.8 and HWE p-value <1E-8, resulting in 5,383,557 remaining SNPs. Ancestry and population structure analyses were performed using the R packages GENESIS [24], SNPRelate [25] and GWASTools [26]. The method was based on the GENESIS vignette on population structure and relatedness [27] (with the exception of square root of 0.05 as the linkage disequilibrium threshold). We estimated the ancestry of the Gen3G participants by jointly analyzing the genetic data from Gen3G and the 1000 Genomes Project, phase 1 [28] (Fig. S1).

### miRNA quantification

Circulating miRNAs from 444 samples were purified as previously described in Légaré et al. [29]. Briefly, total RNA was extracted from 0.5 ml of plasma 1 hour following a 50g glucose challenge test during the first trimester of pregnancy using the mirVana PARIS kit (Thermo Fisher Scientific, catalog # AM1556). Libraries were prepared using the Truseq Small RNA Sample Prep kit (Illumina, BC, Canada; catalog # RS-200-0012) then sequenced on an Illumina HiSeq4000 and HiSeq2500 at the McGill University and Génome Québec Innovation Centre (QC, Canada). Six samples with less than 1 millions high quality reads were excluded, for a total of 369 samples from which genotype data was also available. The extracellular RNA processing toolkit (exceRpt) v4.6.3 [7] was used to process the sequencing data to map high quality reads to the human genome (GRCh37) and miRBase [8] version 21 with STAR [8,30]. From the 2170 miRNAs detected among all samples, 426 mature miRNAs potentially originating from multiple genomic locus were removed in order to specifically identify *cis*-miR-QTLs, leaving 1744 miRNAs. A total of 550 miRNAs with a minimum of 6 mapped reads and 0.1 transcript per million (TPM) in a minimum of 20% of samples were retained for the miR-QTL analysis.

### miR-QTL identification

The miR-QTLs were identified using an expression quantitative trait loci (eQTL) analysis similar to the pipeline applied by the GTEX Consortium (2020) (github.com/broadinstitute/gtex-pipeline/tree/master/qtl). Briefly, the miRNA counts were normalized as with edgeR [31] and the resulting scaled counts per million (CPM) were inverse normal transformed. The mapping window (i.e. the maximum distance for *cis* associations) was set to 1Mb upstream and downstream of the first nucleotide of the mature miRNAs. To assess the associations between the 550 miRNAs and the 5,391,286 SNPs, we used FastQTL [10] with the adaptive permutation mode (1000 to 10,000 permutations) and the following covariates: 6 genotype PCs, sequencing run, gestational age, age of participants, and 25 factors from the Probabilistic Estimation of Expression Residuals (PEER) software [32]. The genotype PCs and PEER factors were included to account for population structure in the Gen3G cohort (as suggested by the scree plot (Fig. S1B)) and for unmeasured variables adding variance in miRNA abundance, respectively. Following the GTEx pipeline, the results from FastQTL were then used to identify the miRNAs with at least one significant association. In more details, a false discovery rate (FDR) threshold of 0.05 was applied on q-values [33] calculated from FastQTL p-values to select 147 miRNAs. For each selected miRNA, significant *cis*-miR-QTLs were then identified using a calculated nominal *p-*value threshold obtained from permutations for that miRNA.

### Construction of genetic risk scores

Using the SNPs identified as miR-QTLs, we built a genetic risk score (GRS) for each miRNA. As previously described [22], SNPs were selected using the elastic net method with the glmnet package [34]. As shown in Equation 1, for each SNP *i* part of a miR-QTL, a GRS assuming an additive genetic effect was calculated for each participant *j* by summing the product of the dosage of effect alleles (G), weighted by its effect size (S) calculated with the elastic-net regression:

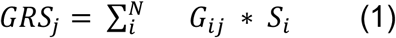

GRSs were calculated for 569 samples since only genotypes are required for the calculation. We used the individual GRS to predict the miRNA abundance from a linear regression model with no covariates.

### Association between GRS and pregnancy complications

To evaluate the association between the predicted miRNA abundance and each outcome, we applied logistic regression between predicted levels and pregnancy complications status for each miRNA. Significant associations with GDM, PE or GH were determined at *p*-value <0.05.

## Data Availability

All data produced in the present study are available upon reasonable request to the authors

## Abbreviations

CI: confidence interval
CPM: count per million
eQTL: expression quantitative trait loci
FDR: false discovery rate
GDM: gestational diabetes mellitus
Gen3G: Genetics of Glucose Regulation in Gestation and Growth
GH: gestational hypertension
GRS: genetic risk score
GTEx: Genotype-Tissue Expression
miRNA: microRNA
miR-QTL: microRNA expression quantitative trait loci
MR: Mendelian randomization
PC: principal component
PE: pre-eclampsia
OR: Odd Ratio
SNP: single-nucleotide polymorphism
TMM: trimmed mean of M-values
TPM: transcript per million TPM

## Declarations

### Ethics approval and consent to participate

This study has been approved by the ethics review board of CIUSSS de l’Estrie-CHUS and all participants provided informed written consent for their inclusion in the study, in agreement with the Declaration of Helsinki.

### Availability of data and materials

The processed data, including the miRNA quantification, are available in Table S2-4. The miRNA-Seq raw data will be available upon request and approval from institutional review boards.

### Competing interests

#### Funding

MG was supported by a doctoral research award from Fonds de la recherche du Québec en santé (FRQS). LB and PEJ are research scholars from the FRQS and members of the CR-CHUS, a FRQS-funded Research Center.

### Authors’ contributions

MFH, LB and PEJ designed the study in collaboration with FW and MG; CL, KT et AAC performed data collection; FW conducted bioinformatic analyses with the help of MG and SC; FW, MG, MFH, LB and PEJ interpreted the results and wrote the manuscript. All authors approved the manuscript.

## Acknowledgements

We first want to thank all the participants of the Gen3G cohort. We are also grateful to the McGill University and Génome Québec Innovation Centre staff for their work in miRNA libraries quality control and sequencing. We also thank Calcul Québec and Compute Canada for their support in this research.

## Notes

### Competing Interest Statement

The authors have declared no competing interest.

### Funding Statement

This study was funded by the "Fonds de recherche du Québec en santé” (FRSQ) and the Canadian Institutes of Health Research (CIHR).

### Author Declarations

Ethics committee/IRB of CIUSSS de l’Estrie-CHUS affiliated with the “Universite de Sherbrooke” gave ethical approval for this work

